# Development and Validation of Comorbidity Index to Predict In-hospital Mortality Risk for Patients with Steven-Johnson Syndrome

**DOI:** 10.1101/2023.05.03.23289447

**Authors:** Renxi Li

## Abstract

**Background:** Stevens-Johnson Syndrome (SJS)/toxic epidermal necrolysis (TEN) is a spectrum of severe drug reactions that has a high in-hospital mortality from 5% to 20%. Early risk prediction for in-hospital mortality in SJS/TEN can be informative for clinical management. This study aimed to develop an in-hospital mortality risk calculator for SJS based on patient comorbidities.

**Methods:** Patients who were diagnosed with SJS/TEN were identified in the National Inpatient Sample database between Q4 2015-2020. SJS patients were randomly sampled into two equal experimental and validation groups. The weight for each comorbidity was determined from a multivariable logistic regression to develop into a single SJS Index.

**Results:** There were 4519 SJS (mortality rate 5.05%), 873 SJS-TEN (mortality rate 13.06%), and 759 TEN (mortality rate 17.00%) patients identified. The SJS Index had good discrimination power (*c*-statistic=0.704, 95% CI=0.656-0.753) and was well-calibrated (Brier score=0.049). SJS Index had similar performance in multivariable regression model with all comorbidities (DeLong p-value=0.621) and validation group (DeLong p-value=0.634). The index had significantly better discriminative power than Elixhauser Comorbidity Index (ECI) (DeLong p-value=0.001). Applying the SJS Index to SJS-TEN patients showed comparable discriminative powers (DeLong test p-value=0.284). In TEN patients, SJS Index remained good calibration (Brier score=0.062) but discriminative power decreased (DeLong p-value=0.005). After adjusting for age, race, and ethnicity, SJS Index had improved performance in all groups.

**Conclusions:** The SJS Index can effectively discriminate and predict in-hospital mortality in SJS and was externally validated in NIS. SJS/TEN often has emergent presentation and immediate intervention is required, this risk score can give an initial assessment of the mortality based on medical records alone and thus offer a quick insight for clinicians for management. In addition, this study demonstrated the possibility of the SJS Index being used as one composite metric to develop a more comprehensive risk-calculator score.

## Introduction

Stevens-Johnson Syndrome (SJS)/toxic epidermal necrolysis (TEN) is a spectrum of severe drug reactions that presents with a rapid progression to full-thickness epidermal necrosis [1]. The severity of epidermal detachment determines the categorization of patients, where SJS indicates involvement of less than 10% of the body surface area (BSA), SJS/TEN overlap involves 10% to 30% BSA, and TEN involves more than 30% BSA. SJS/TEN presents high in-hospital mortality rates. A study based on the National (Nationwide) Inpatient Sample (NIS) database, the largest inpatient database in the US that accounts for 20% of all discharges, showed that the in-hospital mortalities around 5%, 20%, and 15% for SJS, SJS/TEN, and TEN, respectively [2].

Because of the high mortality rate, mortality risk predictors are especially useful in SJS/TEN to provide clinical insights. Severity-of-Illness Score for Toxic Epidermal Necrolysis (SCORTEN) scale, which is composed of age, cancer, heart rate, serum blood urea nitrogen (BUN), detached or compromised body surface, serum bicarbonate, and serum glucose, has been widely used in the clinic to predict mortality in SJS/TEN [3]. However, SCORTEN was originally developed based on a single institutional data in France and there have been raising issues in its application that more recently, a refined ABCD-10 scoring system was developed [1]. The ABCD-10 is composed of age, serum bicarbonate level, cancer, dialysis, and BSA greater than 10% [1].

Since SJS/TEN often has emergent presentation and immediate intervention is required [4]. Thus, quicker references to the predicted mortality would have more usefulness. Parameters from a blood test are required in both SCORTEN and ABCD-10 scores, which can delay the assessment of predicted mortality in SJS/TEN patients for hours. Comorbidities are found to be associated with in-hospital mortality in SJS [5]. This study aimed to develop an in-hospital mortality risk calculator for SJS based on patient comorbidities. This risk score can give an initial assessment of the mortality of SJS patients based on their medical records alone and thus offer a quick insight for clinicians for management.

## Methods

The NIS database is the largest in-patient database in the United States and accounts for 20% of all hospital discharges [6]. Patients who were diagnosed with SJS were identified by the International Classification of Diseases, Tenth Revision, Clinical Modification (ICD-10-CM) code of L51.1 from the last quarter of 2015 and 2020. Moreover, patients diagnosed with SJS-TEN, and TEN were identified by ICD-10-CM of L51.3 and L51.2, respectively. Patient age, demographic information, in-hospital mortality record, and records of comorbidities were extracted from NIS. Of the comorbidities, thirty-eight of them were identified by the Elixhauser Software based on ICD-10-CM codes Comorbidity [7].

Half of the SJS patients were randomly sampled in the experimental group for the development of the mortality index. The remaining half of the patients were designated as the validation group for the purpose of confirming the accuracy of the index. The multivariable logistic regression model to anticipate in-hospital mortality included all 38 Elixhauser comorbidities in the experimental group. The comorbidities examined were acquired immune deficiency syndrome, alcohol abuse, anemias due to other nutritional deficiencies, autoimmune conditions, chronic blood loss anemia (iron deficiency), lymphoma, leukemia, metastatic cancer, in situ solid tumor without metastasis, malignant solid tumor without metastasis, cerebrovascular disease, heart failure, coagulopathy, dementia, depression, diabetes without chronic complications, diabetes with chronic complications, drug abuse, complicated hypertension, uncomplicated hypertension, mild liver disease, moderate to severe liver disease, chronic pulmonary disease, neurological disorders affecting movement, other neurological disorders, seizures and epilepsy, obesity, paralysis, peripheral vascular disease, psychoses, pulmonary circulation disease, moderate renal failure, severe renal failure, hypothyroidism, other thyroid disorders, peptic ulcer disease x bleeding, valvular disease, and weight loss.

A single index, the SJS index, was calculated from the regression coefficient of these comorbidities using Sullivan’s method [8,9]. The multivariable logistic regression furnished parameter estimates of the regression coefficient for each comorbidity, which were utilized to determine the weight for each comorbidity in the mortality index. This was achieved by dividing the regression coefficient of each comorbidity by the absolute value of the smallest regression coefficient and rounding off the quotient to the closest integer. This way, a larger coefficient of comorbidity in the regression was translated into a greater contribution to the mortality index, and vice versa.

The validation group was used to evaluate the predictive ability of the SJS index. Additionally, the experimental groups were used to assess the predictive ability of the Elixhauser Comorbidity Index (ECI) and compared it to that of the SJS index. To demonstrate how the SJS index could be applied, demographic information that is known as risk factors of in-hospital mortality, including age, race, and ethnicity, was incorporated into a multivariable logistic regression analysis to predict in-hospital mortality. Furthermore, the SJS index and demographic-adjusted SJS index were utilized to predict mortality for patients with SJS-TEN and TEN, respectively.

A plot was generated to display the receiver operating characteristic (ROC) curves for each regression model. The AUC, which represents the area under the ROC curve, was computed. To evaluate the ability of each model to distinguish in-hospital mortality, the c-statistic was used, and a 95% confidence interval was determined. A c-statistic of greater than 0.7 indicates that the model has adequate predictive power, while a value between 0.6 and 0.7 suggests moderate predictive power. To compare two different AUCs, a two-tailed DeLong test was used [10].

AUC only assesses the ability of the models to discriminate the occurrence of mortality. However, calibration should also be assessed, since good discrimination may be associated with low calibration that under or over predict mortality [11]. To quantify calibration, Brier score was calculated to measure the difference between the predicted and actual risk [12]. The lower the Brier score is, the closer the predicted and actual risk is, and t^4^hus the better calibrated the SJS index is [12].

SAS (version 9.4) was used to conduct all analyses in this retrospective study, which relied on an open database and therefore did not require IRB approval. The author had complete access to the data and assumes responsibility for the accuracy of the data analysis.

## Results

There were 4519 patients diagnosed with SJS identified in NIS between the last quarter of 2015 and 2020. The total mortality rate was 5.05% (228 cases). In the experimental group, there were 2261 SJS identified with a mortality rate of 5.35% (121 cases). In the validation group, 2258 SJS cases were identified with a mortality rate of 4.74% (107 cases). There were 873 SJS-TEN cases identified with a 13.06% mortality rate (114 cases). For TEN, there were 759 cases identified and the mortality rate was 17.00% (129 cases).

Table 1 presents a summary of the comorbidities and their respective weights used to calculate the SJS index in the experimental group. Only the comorbidities that were presented in the experimental group were included in the calculation, and any comorbidities that were not presented were excluded.

**Table 1.** Comorbidities and their weights in the SJS Index in SJS patients (experimental group) were identified from the NIS database between the last quarter of 2015 and 2020. ***Abbreviations:*** NIS, National (Nationwide) Inpatient Sample; SJS, Steven-Johnson Syndrome.

| **Comorbidities** | **n (%)** | **Weight** |
| --- | --- | --- |
| Acquired immune deficiency syndrome | 36 (1.59%) | 26 |
| Alcohol abuse | 92 (4.07%) | 36 |
| Autoimmune conditions | 202 (8.93%) | -6 |
| Lymphoma | 48 (2.12%) | 43 |
| Leukemia | 34 (1.50%) | 15 |
| Metastatic cancer | 51 (2.26%) | 62 |
| Solid tumor without metastasis, in situ | 2 (0.09%) | -455 |
| Solid tumor without metastasis, malignant | 58 (2.57%) | 41 |
| Cerebrovascular disease | 66 (2.92%) | -13 |
| Dementia | 119 (5.26%) | 3 |
| Depression | 348 (15.39%) | -10 |
| Diabetes without chronic complications | 219 (9.69%) | 11 |
| Diabetes with chronic complications | 406 (17.96%) | 6 |
| Drug abuse | 106 (4.69%) | -60 |
| Hypertension, complicated | 566 (25.03%) | 41 |
| Hypertension, uncomplicated | 622 (27.51%) | -12 |
| Liver disease, moderate to severe | 4 (0.18%) | 47 |
| Chronic pulmonary disease | 508 (22.47%) | 7 |
| Obesity | 403 (17.82%) | 1 |
| Paralysis | 54 (2.39%) | 31 |
| Peripheral vascular disease | 106 (4.69%) | 38 |
| Renal failure, severe | 102 (4.51%) | -26 |
| Hypothyroidism | 314 (13.89%) | -7 |
| Other thyroid disorders | 28 (1.24%) | 47 |
| Valvular disease | 24 (1.06%) | -48 |

In the experimental group of SJS patients, the average SJS index was 11.58 ± 34.20 with a min of -471 and a maximum of 157. In the validation group, the SJS index had an average of 11.98 ± 31.24 with a min of -95, and a maximum of 176. The AUCs of different models in SJS patients were summarized in Table 2 and Figure 1. In the experimental group of SJS patients, in-hospital mortality was adequately discriminated (*c*-statistic = 0.704, 95% CI = 0.656-0.753) and well-calibrated (Brier score = 0.049) by the SJS Index. In the SJS validation group, the ability of the SJS Index to discriminate mortality (*c*-statistic = 0.685, 95% CI = 0.633-0.738) was comparable to that in the SJS experimental group (DeLong test p-value = 0.634). The model in the validation group was also well-calibrated (Brier score = 0.044). The SJS index had similar (DeLong test p-value = 0.621) discrimination power as the multivariable regression model with all comorbidities (*c*-statistic = 0.723, 95% CI = 0.675-0.772). Similarly, the multivariable regression model had a good calibration (Brier score = 0.049). The SJS index has significantly better discriminative power (DeLong test p-value = 0.001) than ECI (*c*-statistic = 0.581, 95% CI = 0.521-0.635), despite ECI had a good calibration (Brier score = 0.050). After adjusting for demographics, the SJS Index had an improved discriminative power (*c*-statistic = 0.752, 95% CI = 0.711-0.793, DeLong test p-value = 0.206) with good calibration (Brier score = 0.048).

**Table 2.** Assessment of discrimination and calibration of in-hospital mortality by SJS Index. SJS, SJS-TEN, and TEN patients were identified in the NIS database between the last quarter of 2015 and 2020. ^*^Reference group is SJS Index, SJS-TEN. ^**^Reference group is SJS Index, TEN. ***Abbreviations:*** NIS, National (Nationwide) Inpatient Sample; ROC, receiver operating characteristic; SJS, Steven-Johnson Syndrome; TEN, toxic epidermal necrolysis.

| Predictive Model | c-statistic (95% CI) | Brier Score | DeLong P-value |
| --- | --- | --- | --- |
| SJS Index, Experimental Group | 0.704 (0.656-0.753) | 0.049 | Ref. |
| SJS Index, Validation Group | 0.685 (0.633-0.738) | 0.044 | 0.634 |
| All Comorbidities | 0.723 (0.675-0.772) | 0.049 | 0.621 |
| ECI | 0.581 (0.521-0.635) | 0.050 | 0.001 |
| Demographics-adjusted SJS Index | 0.752 (0.711-0.793) | 0.048 | 0.206 |
| SJS Index, SJS-TEN | 0.661 (0.611-0.711) | 0.051 | 0.284 |
| SJS Index, TEN | 0.594 (0.541-0.648) | 0.062 | 0.005 |
| Demographics-adjusted SJS Index, SJS-TEN | 0.726 (0.682-0.771) | 0.107 | 0.111* |
| Demographics-adjusted SJS Index, TEN | 0.626 (0.573-0.679) | 0.137 | 0.428** |

**Figure 1.**
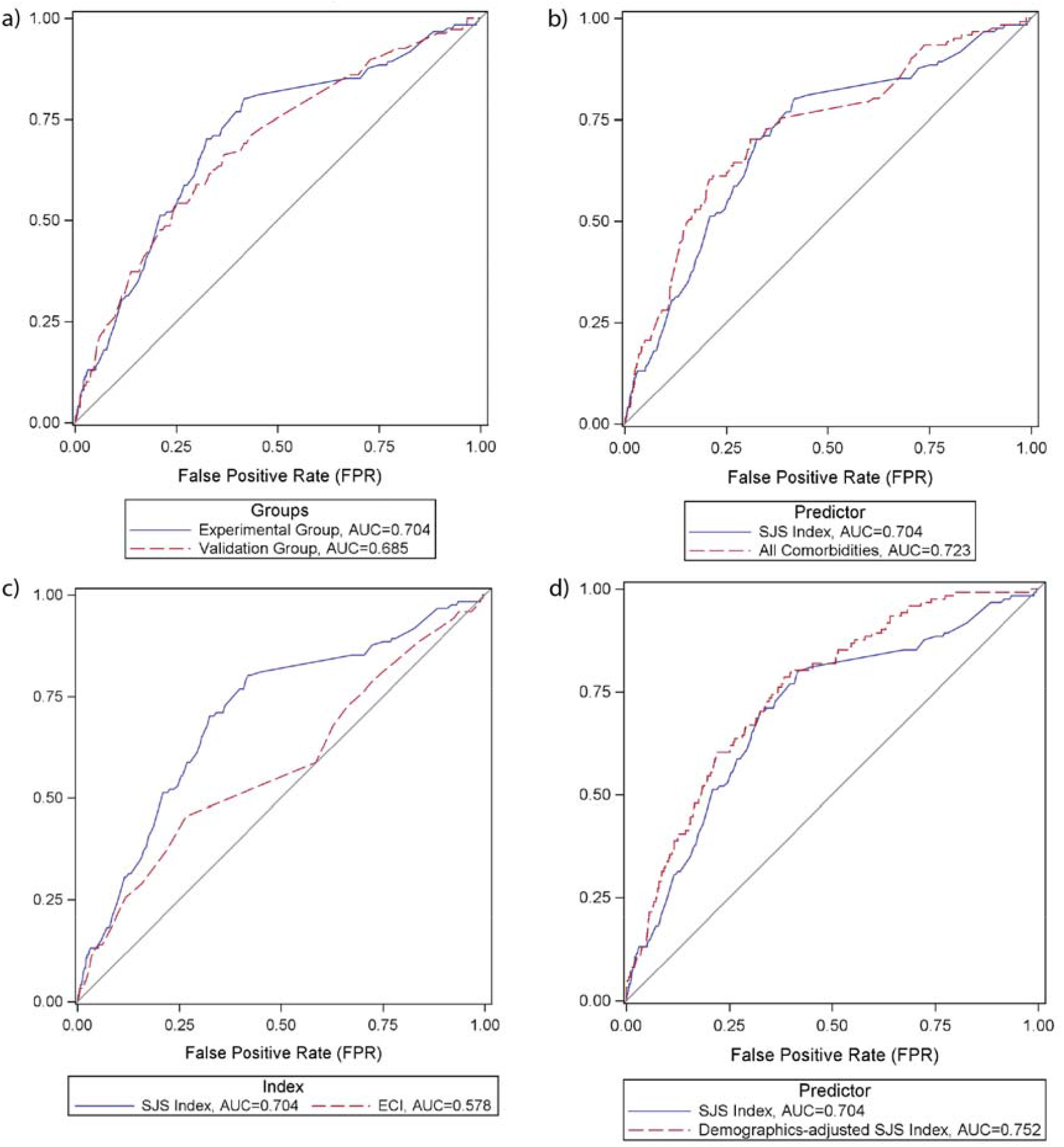
The ROC performance in predicting in-hospital mortality for SJS patients from Q4 of 2015 to 2020 in NIS was compared in four scenarios. a) Comparison of the SJS Index between the experimental and validation groups. b) Comparison of the SJS Index with a multivariable regression model that included all comorbidities in the experimental group. c) Comparison of the SJS Index with the ECI in the experimental group. d) Comparison of the SJS Index with the demographics-adjusted SJS Index in the experimental group. ***Abbreviations:*** NIS, National (Nationwide) Inpatient Sample; ROC, receiver operating characteristic; SJS, Steven-Johnson Syndrome.

The SJS Index were applied in SJS-TEN and TEN patients to predict their in-hospital mortality and the AUCs of different models were summarized in Table 2 and Figure 2. The discrimination power of the SJS Index was moderate in SJS-TEN patients (*c*-statistic = 0.661, 95% CI = 0.611-0.711) and was not significantly different from that in SJS patients (DeLong test p-value = 0.284). The model in SJS-TEN patients was well-calibrated (Brier score = 0.051). SJS Index, however, performed significantly inferior in terms of discrimination in TEN patients (*c*-statistic

**Figure 2.**
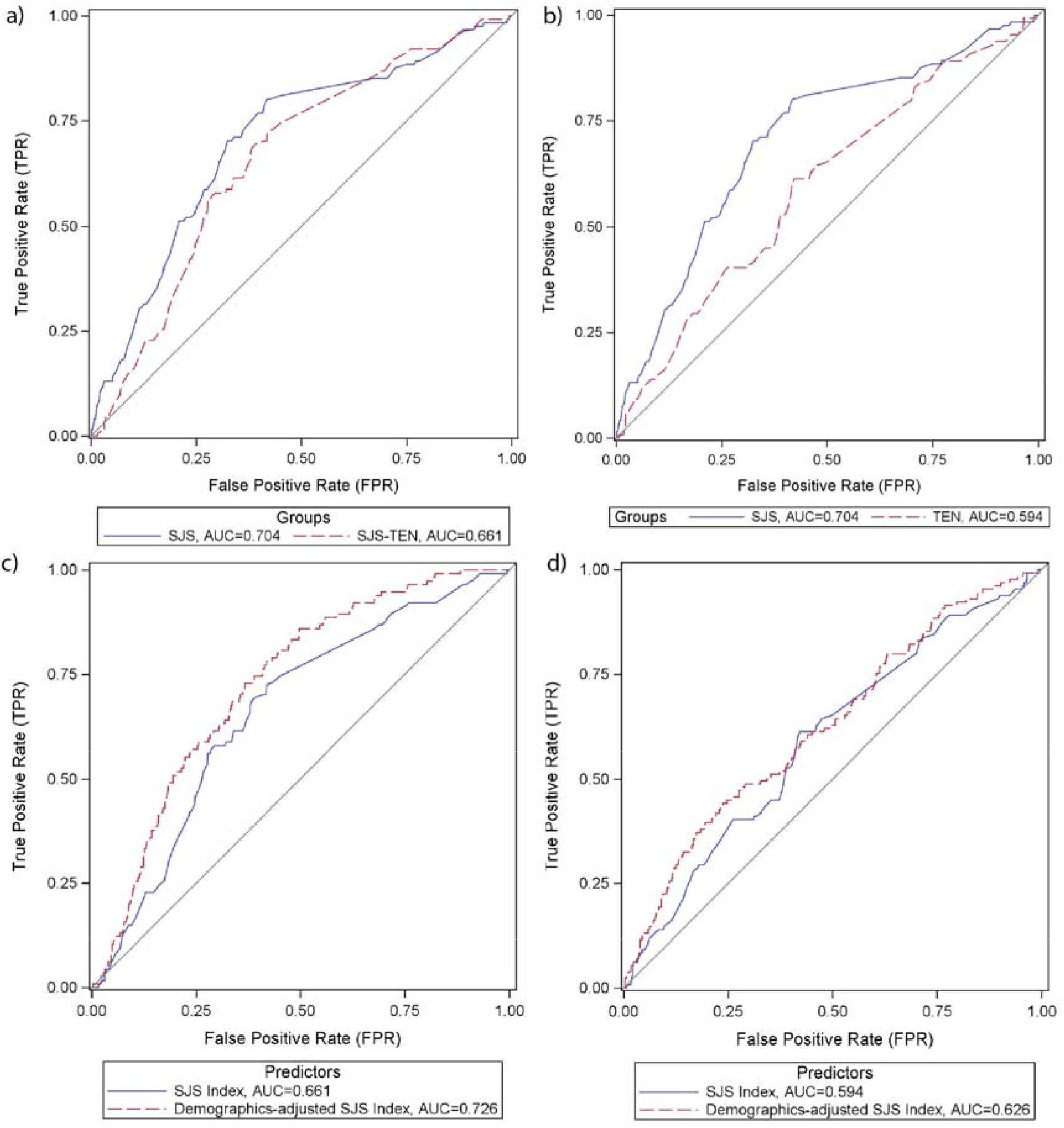
The ROC performance in predicting in-hospital mortality for SJS, SJS-TEN, and TEN patients from Q4 of 2015 to 2020 in NIS was compared in four scenarios. a) Comparison of the SJS Index between SJS (experimental group) and SJS-TEN patients. b) Comparison of the SJS Index between SJS (experimental group) and TEN patients. c) Comparison between SJS Index and demographics-adjusted SJS Index in SJS-TEN patients. d) Comparison between SJS Index and demographics-adjusted SJS Index in TEN patients. ***Abbreviations:*** NIS, National (Nationwide) Inpatient Sample; ROC, receiver operating characteristic; SJS, Steven-Johnson Syndrome; TEN, toxic epidermal necrolysis.

= 0.594, 95% CI = 0.541-0.648, DeLong test p-value = 0.005) despite a good calibration (Brier score = 0.062). After adjusting for demographics, SJS Index adequately discriminated in-hospital mortality in SJS-TEN patients (*c*-statistic = 0.726, 95% CI = 0.682-0.771) while the calibration went down (Brier score = 0.107). In TEN patients, demographics adjusted SJS Index moderately discriminated in-hospital mortality (*c*-statistic = 0.626, 95% CI = 0.573-0.679) and calibration similarly went down (Brier score = 0.137).

The correspondence between the SJS score (range = -100 to 500) and predicted in-hospital mortality is shown in Figure 3 and the selected correspondence was listed in Table 3. The in-hospital mortality can be calculated as follows: 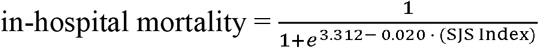.

**Figure 3.**
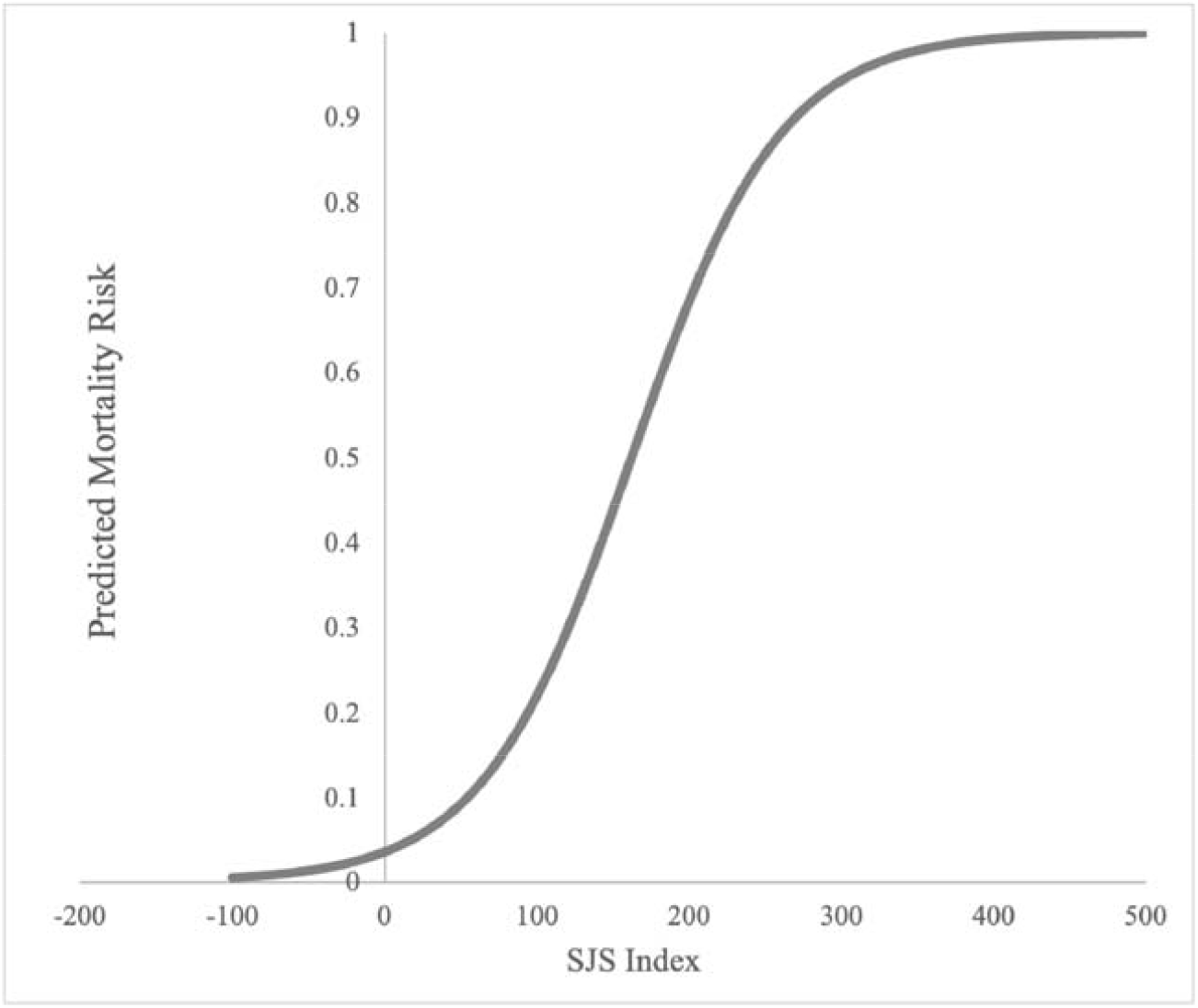
The logistic curve of the in-hospital mortality risk predicted by the SJS Index in SJS patients (development group) from Q4 of 2015 to 2020 in NIS. ***Abbreviations:*** NIS, National (Nationwide) Inpatient Sample; SJS, Steven-Johnson Syndrome.

**Table 3.** Corresponding predicted in-hospital mortality risk to SJS Index derived from the logistic curve in SJS patients (development group) from Q4 of 2015 to 2020 in NIS. ***Abbreviations:*** NIS, National (Nationwide) Inpatient Sample; SJS, Steven-Johnson Syndrome.

| SJS Index | Predicted In-hospital Mortality (%) |
| --- | --- |
| -100 | $0.47 \pm 0.14$ |
| -50 | $1.30 \pm 0.24$ |
| 0 | $3.52 \pm 0.43$ |
| 50 | $9.18 \pm 1.54$ |
| 100 | $21.89 \pm 5.05$ |
| 150 | $43.74 \pm 10.34$ |
| 200 | $68.31 \pm 11.88$ |
| 250 | $85.67 \pm 8.34$ |
| 300 | $94.31 \pm 4.35$ |
| 350 | $97.87 \pm 1.96$ |
| 400 | $99.22 \pm 0.83$ |
| 450 | $99.72 \pm 0.34$ |
| 500 | $99.90 \pm 0.14$ |

## Discussion

This study identified SJS patients in the NIS database from the last quarter of 2015 to 2020 and developed an in-hospital mortality risk score for SJS based on patient comorbidities.

The multivariable regression model accounting for all comorbidities identified by Elixhauser measure adequately discriminated and predicted (*c*-statistic = 0.723, 95% CI = 0.675-0.772, Brier score = 0.049) in-hospital mortality in SJS patients. Sullivan’s method successfully translate the multivariable regression model into the single SJS index, indicated by a non-significant DeLong test p-value = 0.621 for AUC [8,9]. The discriminative power of the SJS index did not differ (DeLong test p-value = 0.634) between the experimental group and the independent SJS patient sample (validation group). This demonstrated the external validity of the SJS Index in the NIS database and thus the success of the index development. In future studies, additional data sources should be used to validate the SJS Index.

ECI is a weighted single-scoring system to predict in-hospital mortality [13]. However, ECI was developed based on Elixhauser Comorbidity in all patient populations [13]. ECI had high discriminative power (*c*-statistic = 0.777, 95% CI = 0.776-0.778) in the validation phase of its development in all patients [13]. Although ECI has been validated in a number of high-mortality disease categories such as septicemia and pneumonia, it has not been assessed in SJS/TEN patient population [13]. This study examined ECI and found ECI did not have adequate discriminative power (*c*-statistic = 0.581, 95% CI = 0.521-0.635) despite a good calibration (Brier score = 0.050). The discriminative power of the SJS Index was significantly greater than that of ECI (DeLong test p-value = 0.001). This demonstrated the comorbidity index developed from a non-specific patient population could not be sufficiently applied in SJS patients and the development of the SJS-specific comorbidity index in this study was necessary.

Applying the SJS Index to SJS-TEN patients showed comparable discriminative powers (DeLong test p-value = 0.284) and good predictive power (Brier score = 0.051) as in SJS patients. In TEN patients, the predictive power of the SJS Index remained good (Brier score = 0.062) but the discriminative power of the SJS Index significantly decreased (DeLong test p-value = 0.005) compared to the SJS patients. This demonstrated although developed in SJS patients only, the SJS Index was also highly effective in SJS-TEN patients. This could be attributed to overlapping characteristics between SJS and SJS-TEN patients. In TEN patients, however, the SJS Index was less effective. TEN patients have more severe clinical presentations by the drug reaction. Thus, the state of the patients at presentation may be a greater factor for mortality than comorbidities.

Besides using as a single index to give an initial assessment of predicted in-hospital mortality in SJS/TEN patients, this study also showed the possibility of using the SJS Index in combination with other metrics for a more accurate assessment of mortality. The study included the SJS Index and relevant demographic information, including age, race, and ethnicity, in a multivariable regression and estimated the discriminative powers of the model. Although not significant, AUC for discriminating in-hospital mortality increased from 0.704 to 0.752 in SJS (DeLong test p-value = 0.206), from 0.661 to 0.726 in SJS-TEN (DeLong test p-value = 0.111), and from 0.594 to 0.626 in TEN (DeLong test p-value = 0.428). NIS does not record relevant clinical information such as lab values and BSA. However, in other datasets, the SJS Index can be used in combination with patients’ clinical information to develop a more comprehensive scoring system. This will be superior to the SCORTEN and ABCD-10 scores in the way that comorbidities in the new scoring system will be more comprehensive.

There were several limitations of this study. SJS/TEN patients continue to have a high risk for mortality after discharge [14]. However, NIS only records in-hospital mortality without any follow-up. Thus, post-discharge mortality cannot be accounted for when developing or validating the SJS Index. In further studies, the ability of the SJS Index to predict post-discharge mortality should be examined in other datasets. Secondly, the comorbidities were based on the Elixhauser measure, which recorded binary (yes/no) for each comorbidity. As a result, the comorbidity did not account for the extent and/or staging of the disease, such as the class of obesity. Binarization of the comorbidities will make it easier to record and calculate the SJS Index but providers should be aware of the heterogeneity within the comorbidity.

In this study, SJS patients were identified in the NIS database from Q4 of 2015 to 2020, and a risk score for in-hospital mortality in SJS patients was created using patient comorbidities. The SJS Index had good discrimination power and was well-calibrated. The index was externally validated in the NIS database and performed significantly better than the non-specific ECI. The SJS Index was effective in SJS-TEN patients but had decreased effectiveness in TEN patients. When combined with relevant demographic information, the SJS Index showed better performance, demonstrating the possibility of the SJS Index being used as one composite metric to develop a more comprehensive risk-calculator score. The correspondence between the SJS score and predicted in-hospital mortality was charted and plotted for quicker reference in the clinics.

## Data Availability

All data produced in the present study are available upon reasonable request to the authors

## Statements and Declarations

### Conflict of Interest

Renxi Li declares no conflict of interest.

Informed Patient Consent: Given the use of retrospective, de-identified NIS data, this study was exempted from IRB approval.

## Acknowledgments

Renxi Li contributed to all aspects of this study. The author acknowledges the guidance and support of colleagues and reviewers who provided useful feedback throughout the work.

